# Feasibility and preliminary efficacy of an online mindful walking intervention among COVID-19 long haulers: A mixed method study including daily diary surveys

**DOI:** 10.1101/2024.01.23.24301694

**Authors:** Abhishek Aggarwal, Shan Qiao, Chih-Hsiang Yang, Slone Taylor, Cheuk Chi Tam, Xiaoming Li

## Abstract

**Objective:** Mindful-walking (MW) intervention could be an effective strategy to address the psychosocial stressors and physical health challenges faced by COVID-19 long haulers. This study aims to test the feasibility of digitally delivering MW intervention among long haulers via social media and assess its preliminary efficacy on enhancing physical and psychosocial wellbeing.

**Method:** We recruited 23 participants via Facebook groups in March and November 2021 for a 4-week online MW intervention, consisting of mindfulness practices (2 sessions per week), delivered entirely through the Facebook group. The intervention was assessed using mixed methods. Quantitative data were collected through 28-day brief daily evening surveys over the 4 weeks intervention period, including affect, cognition, mindfulness, physical activity, and MW engagement. Qualitative data were extracted from Paradata (i.e., participants’ responses to the social media posts). Multilevel modeling was employed for statistical analysis and a pragmatic approach was used for qualitative analysis.

**Results:** The mean feasibility score was 4.93/7 (*SD=*1.88). Multilevel models showed that MW uptake on a given day was positively associated with positive affect (*β*=0.89, *p*<0.01), perceived cognition (*β*=0.52, *p*<0.05), and physical activity levels (*β*=0.41, *p*<0.05), and negatively associated with negative affect (*β*=-0.83, *p*<0.01). Total number of MW days across the study period were positively associated with mindfulness levels (*β*=0.3 *p*<0.01). Paradata reported satisfaction in mindfulness skill enhancement, symptom management and well-being promotion.

**Conclusion:** The digital delivery of our MW intervention via Facebook showed high acceptability. Preliminary efficacy findings indicate improved mental wellbeing and physical activity among long haulers.

## Introduction

According to the World Health Organization (WHO), as of October 2023, the novel coronavirus disease 2019 (COVID-19) infected over 771.5 million individuals and claimed over 7 million lives (WHO, 2023). To control the COVID-19 pandemic, several restrictions were imposed, including wearing masks, maintaining social distance, travel bans, and isolation (Baloch et al., 2020; CDC, 2020, 2022a; World Health Organization, 2021). About 30% of COVID patients, termed “long-haulers,” experienced persistent symptoms, even up to twelve weeks post-infection, that cannot be explained by an alternative diagnosis such as chronic fatigue, cognitive issues, and loss of taste and smell (Bull-Otterson, 2022; CDC, 2022b; Wong et al., 2021). These symptoms are associated with severe health challenges for long haulers, including: (1) cognitive and psychiatric concerns, such as Post-Traumatic Stress Disorder (PTSD), cognitive impairments, attention deficits, and depression (Graham et al., 2021; Ladds et al., 2020); and (2) physical ailments, including exhaustion, muscular pain, and breathing difficulties (H. E. Davis et al., 2021). Additionally, long haulers also confront socio-economic setbacks, including social isolation, job loss, and reduced earnings (Aghaei, Zhang, et al., 2022; Okoloba et al., 2020; Otache, 2020). Notably, existing data have shown that many long haulers turn to substance misuse for managing their cognitive and psychiatric problems (Tam et al., 2023), increasing the risk for additional health issues, such as drug overdose. Given these multifaceted challenges, a psycho-behavioral health promotion intervention addressing the quality of life in long haulers becomes warranted.

Outdoor physical activity holds strong potential as an effective coping and preventive strategy given its many well-documented physical, social, and mental health benefits for people of all ages, especially those with or at risk of developing chronic diseases (Reiner et al., 2013). A recent cross-sectional study reported that physical activity during the pandemic was associated with a reduced likelihood of long COVID and a reduced duration of long COVID symptoms, including fatigue, neurological complications, cough, and loss of sense of smell or taste (Feter et al., 2023). Therefore, physical activity can play a crucial role in the recovery of long haulers, including regaining their physical stamina post-illness (di Fronso et al., 2022). However, COVID-19 negatively impacted physical activity levels due to lockdowns, isolation, and health issues (Caputo & Reichert, 2020; Grocke-Dewey et al., 2021; Watson et al., 2021; Wilke et al., 2022). Especially, long haulers have reported lower physical activity levels, and increased fatigue severity (Vélez-Santamaría et al., 2023). A recent rapid review of 61 studies from Asian and European countries, with a small portion from the US, found that the pandemic led to significant reductions in mobility, walking, and physical activity, along with an increase in sedentary behavior (Park et al., 2022). There is a clear need for health promotion interventions with long haulers to incorporate physical activity.

Mindfulness practice may be a promising component of physical activity interventions due to its emphasis on connecting the physical and sensational cues with mental health promotion. Unlike traditional therapies such as Cognitive Behavioral Therapy (CBT), mindfulness is preventive, self-initiated, easily integrated into daily routines, and less resource-intensive while offering comparable benefits (J. Li et al., 2021). This approach has been shown to effectively reduce anxiety, depression, and distress in patients with chronic diseases, including cancer and diabetes, as highlighted in several systematic reviews (Guo et al., 2019; P. Li et al., 2022; Oberoi et al., 2020). Furthermore, mindfulness has the potential to enhance physical activity levels by influencing its psychological determinants. Studies have demonstrated that mindfulness can positively impact physical activity by fostering greater awareness and attention to present experiences, thereby promoting a more active lifestyle (Schneider et al., 2019; Yang & Conroy, 2020).

Mindfulness involves heightened attention and awareness of the present moment, as well as the acknowledgement of one’s current physical and emotional sensations (Bishop et al., 2004; Brown & Ryan, 2003). It can be conceptualized as both a “trait” and “state”. Trait mindfulness refers to an individual’s dispositional characteristic of mindfulness that varies between individuals, while state mindfulness refers to momentary levels of experienced mindfulness, influenced by situational factors, that varies within individuals (Brown & Cordon, 2009; Yang & Conroy, 2020). Many mindfulness-based programs, such as Mindfulness-Based Stress Reduction (MBSR), primarily emphasize static meditation practices, which may not cater to the physical activity needs of individuals, such as long haulers who need to regain physical stamina post-illness (di Fronso et al., 2022). Integrating mindfulness techniques with routine physical activities, such as walking, can enhance state mindfulness while promoting physical activity (Aghaei, Aggarwal, et al., 2022). Mindful walking (MW), a low-intensity, accessible exercise, could help in gently rebuilding physical strength and endurance for long haulers, without overly exerting themselves, and assist in managing anxiety or stress associated with their recovery process (Lin & Yeh, 2021; Sun et al., 2022; Teut et al., 2013). Hence, MW may foster both physical and mental rehabilitation for long haulers, aligning with their health goals, while also easily fitting into daily routine life for sustained practice.

A growing number of interventions have been developed to address the well-being of long haulers. A comprehensive scoping review highlighted a range of interventions from singular approaches like pharmacological treatments, electromagnetic field sessions, and dietary modifications, to holistic programs encompassing multiple health components (Al-Jabr et al., 2023). These integrated programs touched upon aspects such as rehabilitation, lifestyle changes, stress management, sleep hygiene, breathing techniques, dietary optimization, energy conservation, psychoeducation, maintaining social interactions, and cognitive behavioral therapy (CBT). While some interventions emphasized either mindfulness practice or physical activity, few combined both (D. Davis et al., 2022). Additionally, none of the MW interventions targeted long haulers. This observation underscores a clear gap in the literature and the importance of evaluating the feasibility and preliminary efficacy of a MW program with long haulers.

In context of the COVID-19 pandemic, the adoption of online platforms for healthcare interventions has become indispensable. Social media or online social networking sites (for e.g., such as Facebook, Twitter, Reddit, Pinterest, and Instagram) provide a means for users to engage with each other and share content, presenting an opportunity to use this modality for behavior change interventions. Social media usage has been stable over the past 5 years, with 7 in 10 US adults using any kind of social media platform (Anderson, 2021). Particularly, Facebook remain a dominate platform after YouTube with 69% of users reporting its use (Anderson, 2021). The use of social media has evolved, with now more users seeking and exchanging health-related information (Fox, 2014). The pandemic’s physical distancing mandates, transportation barriers, and the inaccessibility to traditional healthcare facilities (Smith et al., 2020) have underscored the importance of such online platforms for interventions. Delivering behavioral interventions via a social media platform eliminates the need for physical visits, offers flexibility in participation timing, integrates interventions seamlessly into daily routines, and offers access to the social resources (Pagoto et al., 2016).

Online or web-based mindfulness interventions have been effective in decreasing depression, anxiety, stress, and improving mindfulness among patients with physical health conditions, including cancer, chronic pain, heart disease, etc. (Z. Liu et al., 2022; Toivonen et al., 2017); however, to date, no studies have explored the feasibility and preliminary efficacy of delivering a MW intervention for long haulers through social media (Yang & Conroy, 2018). A prior pilot study with college students in China demonstrated the potential of a MW intervention delivered via a smartphone app, showing initial effectiveness in improving mental health (Sun et al., 2022).

### The present study

In our study, we aim to pilot-test a MW program delivered through a social media platform for long haulers. Our approach includes administering daily diary surveys to measure participants’ day-level psychological and physical activity outcomes, one-month follow-up survey post-intervention to assess the feasibility of digitally delivering the MW intervention, and qualitative data, gathered online, for a richer understanding of the participants’ personal experiences and feedback. This mixed-method approach is designed not only to evaluate the program’s immediate effects but also to inform refinements for future iterations, ensuring sustained engagement and maximal benefits for long haulers.

## Methods

### Recruitment

Participant recruitment was conducted over 7 months from the end of March 2021 to the end of October 2021 through purposive and snowball sampling, primarily using social media platforms such as Facebook, Slack, Reddit, etc. In total, 16 Facebook groups, 1 Slack group, 1 Reddit subthread, and 2 organization websites were identified targeting COVID-19 and/or COVID-19 long haulers. Upon receiving approval from administrators of the groups and organizations, recruitment posts were added to 8 Facebook groups, 1 Slack group, 1 organization website, and 1 Reddit subthread. Six out of the 8 Facebook groups and the Slack group were private, meaning only those who were members of the group could see the post. The number of members in the Facebook groups ranged from 1,000 to 166,000 members.

### Inclusion and Exclusion Criteria

In total, 46 individuals contacted the researcher through email or Facebook Messenger to participate in the study. They were eligible for the study if they 1) were 18 years or older, 2) could speak and understand English, 3) had been infected with COVID-19, 4) had experienced at least one COVID-19 symptom for 4 weeks or longer after COVID-19 diagnosis, and 5) had no issues walking at a normal pace. Out of 44 people met the inclusion criteria, and 23 people participated in the 4-week intervention (Figure 1), remaining participants reported either a lack of interest or physical inability to participate.

**Figure 1.**
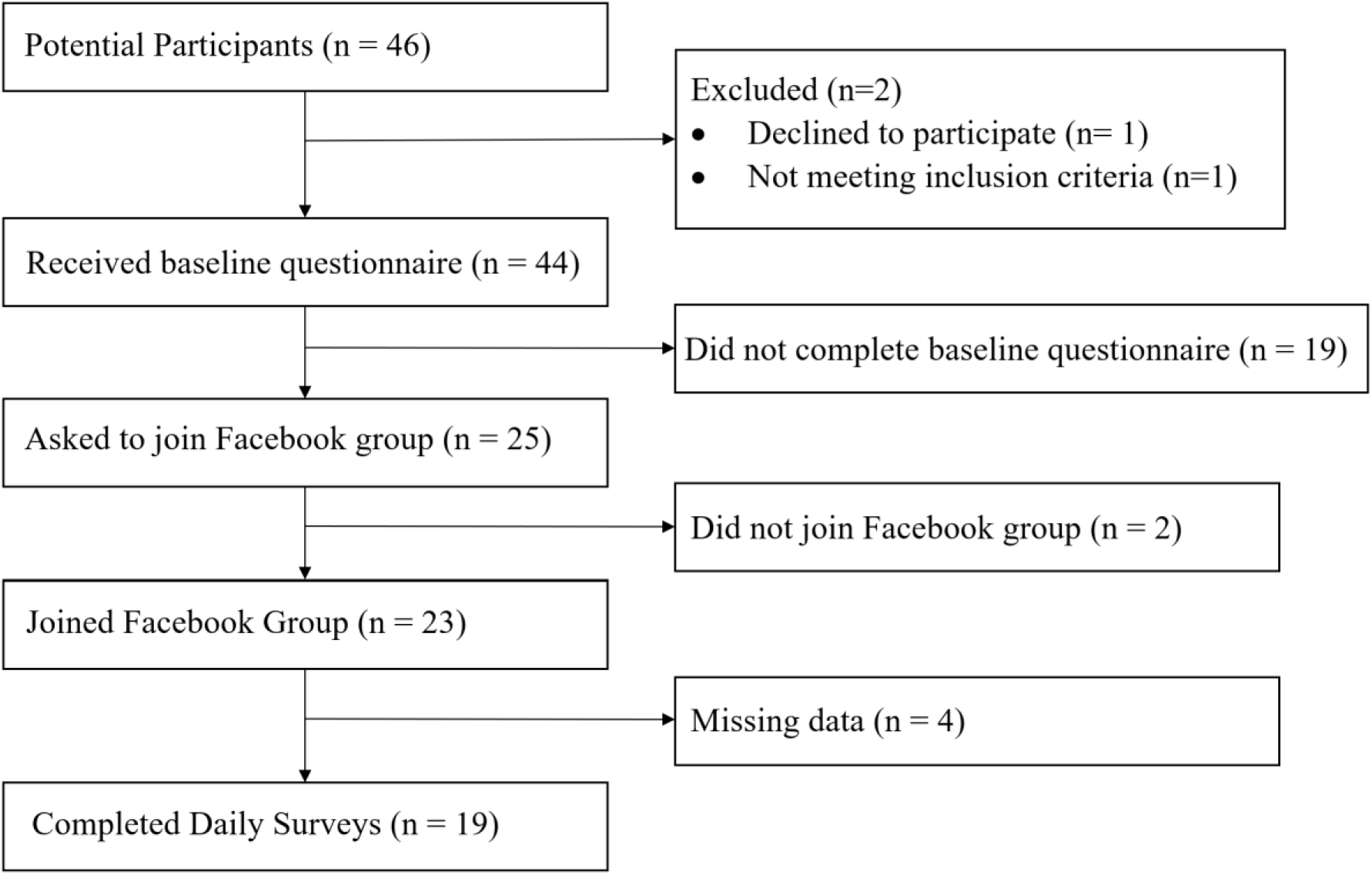
Participant screening

### Mindfulness Walking Intervention

The MW sessions, adapted from (Yang & Conroy, 2020), are modified from the conventional mindfulness meditation practices (Kabat-Zinn, 1994). The sessions constituted three primary practices: mindful breathing walking, step-focused walking, and full body scan walking. Mindful breathing focuses on paying attention to the rhythm of the breath i.e., each inhale and exhale. Step-focused walking focuses on paying attention to the heel-to-toe rhythm as each foot makes contact with the ground. Full body scan focuses on elevating attention and awareness of each body part (top-down and bottom-up) to observe any emotional or physical sensations that may arise.

All sessions consisted of slow walking at a pace of approximately one step per second for 30 minutes at a convenient location in the participants’ neighborhood. Each successive week, the three mindfulness practices were progressively introduced and added to the slow walking session. The first session (week 1) did not include mindfulness practice to allow the participants to practice slow walking for 30 minutes. The second session (week 1) included mindful breathing for only 5 minutes of the 30-minute slow walk. The third and fourth sessions (week 2) included mindful breathing for 10 minutes of the 30-minute slow walk. The fifth and sixth sessions (week 3) included 10 minutes of mindful breathing, followed by 10 minutes of step-focused walking during the 30 minutes slow walk. In addition to the mindfulness practices in the fifth and sixth sessions, the seventh and the eighth sessions (week 4) included 10 minutes of a full body scan during the last 10 minutes of the 30-minute slow walk.

### Intervention strategies and procedures

The intervention was implemented for two cohorts, wave 1 (n=12) and wave 2 (n=11). Wave 1 began in August 2021, and wave 2 began in November 2021. Participants were sent a baseline survey via email two weeks prior to the start of the intervention, and those who completed the survey were invited to join a private Facebook group. There were two separate Facebook groups for Wave 1 and Wave 2. Prior to the start of the MW sessions, participants were instructed to watch a pre-recorded orientation video.

Every Monday, a pre-recorded video providing step-by-step instructions on that week’s MW session was uploaded. Participants could complete the week’s session any time during the week but a one-day gap between two sessions within a week was required. The intervention facilitator posted prompt questions to the group after each session to encourage the participants to share their experiences, questions, and feedback. The facilitators checked and replied to the questions and/or responded to participants’ comments every evening.

Participants were also instructed to complete a daily survey via RedCap every evening during the 4-week intervention. The survey was sent out every day over the 4 weeks at 8pm ET via email. The survey focused on participants’ emotional, mental, and physical well-being and their perceived mindfulness in that moment. In the one-month follow-up, the overall feasibility of the intervention was assessed using the Usefulness, Satisfaction and Ease of Use scale (USE) (Lund, 2001). Participants who completed the intervention had a chance to win one of the six $30 e-gift cards.

### Key measures in each type of survey

#### Baseline survey (time-invariant covariates)

##### Demographics

The baseline survey included items on age, gender, race/ethnicity, education level, state and county of residence.

##### COVID-19 related

COVID-19 diagnosis date, ever been hospitalized due to COVID-19, and pre-existing conditions before COVID-19 diagnosis. Another variable i.e., months since COVID-19 diagnosis was calculated by subtracting the COVID-19 diagnosis date from the date on which the demographics survey was filled.

#### One-month follow-up survey

##### USE scale

The overall feasibility or the usefulness, satisfaction, and ease of use of the intervention were assessed using the USE scale at the one-month follow-up (Lund, 2001). The 30-item scale with a 7-point Likert scale ranging from strongly disagree to strongly agree measured usefulness (8 items), ease of use (11-items), ease of learning (4 items), and satisfaction (7 items). An overall mean score of 4 or more was regarded as feasible.

##### Qualitative data

Linguistic data on participants’ interactions with the intervention modules (e.g., skill-learning video, reflection questions, and discussion polls) were collected (Couper, 2017). Data included comments, questions, views, and replies on specific posts from the Facebook group page during the intervention. They were used for the qualitative analysis to assess the acceptability and preliminary efficacy of the intervention program. Responses from the intervention facilitators were excluded from the analyses.

#### Daily evening survey (time-varying covariates)

##### Positive Affect

Positive affect was measured through three items: calm (“How calm/relaxed are you today?”), energetic (“How energetic/excited are you today?”), and happy (“How happy/joyful are you today?”). The response options ranged from 1=Not at all to 7=Extremely. The sum score of these three items represented positive affect. These items were adapted from the “Positive Affect” measure in Patient Reported Outcomes Measurement Information System (PROMIS).

##### Negative Affect

Negative affect was measured through three items: stressed (“How stressed are you today?”), lonely (“How alone/lonely are you today?”), and fatigue (“How fatigue/tired are you today?”). The response options ranged from 1=Not at all to 7=Extremely. The sum score of these three items represented negative affect. These items were adapted from the “Negative Affect” measure in PROMIS (Pilkonis et al., 2011; Schalet et al., 2016).

##### Perceived Cognition

Perceived cognition was measured through two items: sharp (“How sharp is your mind today?”), and concentrate (“I can concentrate and keep track of things today”). The response options ranged from 1=Not at all to 7=Extremely. The sum score of these two items represented perceived cognition. These two items were adapted from the PROMIS Cognitive Function - Abilities Subset (v2.0) to assess perceived cognition in real-time (Fries et al., 2005; Yang et al., 2021).

##### Mindfulness

Mindfulness was measured through two items from the State Mindfulness Scale (SMS, Tanay & Bernstein, 2013). SMS was originally developed to test situational mindfulness i.e., mindfulness levels at a specific time and within a specific context. The overall SMS scale had sound internal consistency (α = .95), along with its two factors: Mindfulness of Body factor (α = .95), and Mindfulness of Mind (α = .90). The two items were selected to represent the two factors. Item 1, emotional awareness (“I am aware of any thoughts or emotions in my today.”), belonged to factor the ‘Mindfulness of Mind’, and item 2, physical awareness (“I am aware of any physical feelings or sensations from my body today”), belonged to factor the ‘Mindfulness of Body’. Both items had 7-point Likert scale response options ranging from 1=Not at all to 7=Extremely. The sum score of both items was used to represent mindfulness.

##### Physical activity minutes on a given day

The physical activity levels of the participants on a specific day were measured through a single item, “Approximately, how much time did you exercise today (not including the MW time)?”.

##### MW on a given day

The participants’ self-reported whether they practiced MW on a given day through a single item, “Did you practice mindful walk today?”. The participants responded as “Yes” or “No” for this item.

##### Days of practicing MW

The total number of days a participant practiced MW was computed by adding up the “Yes” response on “Did you practice MW today?” for each participant separately.

##### Time-varying covariates at the day level

Two time-varying covariates were assessed in the daily surveys, including typical day (“Do you feel sick or ill today?” (Yes/No)), and social engagement (“How many people did you talk to/interact with today?”). Also, “Time of finishing the survey,” i.e., the time at which participants’ entered their daily survey responses was automatically recorded by RedCap.

### Data preparation and analysis

#### Qualitative data analysis

Paradata was analyzed using a pragmatic approach, which has been widely used in mixed method studies (Vaismoradi et al., 2013). This was performed via several steps: (1) extracting data from Facebook group page and familiarizing the data; (2) generating a coding scheme; (3) structuring the data and identifying emergent themes; (4) reviewing coded data and comparing patterns; (5) defining themes and interpretation (Vaismoradi et al., 2013). Descriptive statistics, such as frequencies and percentages of the posts corresponding to a theme, were also calculated.

#### Quantitative data analysis

Mean scores were calculated for the USE scale to evaluate the feasibility of the MW intervention. Multilevel models were used to analyze the data from daily evening survey due to the nested structure of the data (days nested within people). The data cleaning involved two important steps. First, if any participant answered a survey multiple times within a day, then only the last response by the participant was considered. Each participant had only one response per day. Second, if any participant answered the daily prompt post-midnight, but before sleep time (i.e., any time before 3 AM), then that response was attributed to the previous day. Additionally, the distribution of each outcome variable was examined, including positive affect, negative affect, perceived cognition, mindfulness, and physical activity. All variables except physical activity had a reasonable skewness and kurtosis between +0.5 and -0.5 (Ryu, 2011). Physical activity had a skewness and kurtosis of 2.61 and 10.53, respectively. Therefore, the Box-Cox transformation was applied on the outcome physical activity minutes on a given day (Osborne, 2019), which significantly reduced the skewness and kurtosis to 0.11 and -1.83. The transformed variable was used for analysis. Continuous covariates, including age and time of finishing the survey were grand mean centered to avoid multi-collinearity issues and improve interpretability (Bauer & Curran, 2005).

Five multi-level models examined the association of outcome variables on a given day, including positive affect, negative affect, perceived cognition, mindfulness, and physical activity, and their primary predictors i.e., MW on a given day (yes/no), and the total number of MW days across the study, after controlling for demographics and time-varying covariates. Each model included random intercept effects for a better model fit. The models were estimated using R 4.2.0 with the nlme package (r José Pinheiro, et al., 2023). The final multilevel model was:

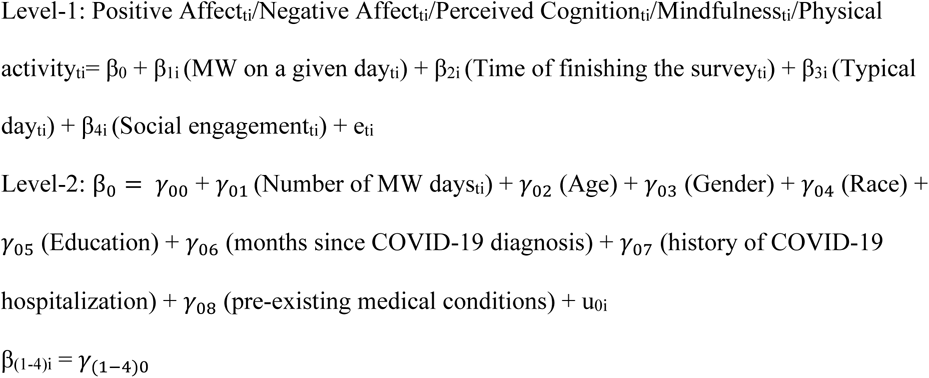

where the level-1 equation constitutes time-varying covariates i.e., variables that change at the daily level, and the level-2 equation constitutes the time-invariant covariates i.e., variables that do not change daily for a person. At level-1 equation, β_0_ is the intercept (defined at level-2), β_(1-4)i_ are the estimate of slope of relationship between outcomes and time-varying covariates, and e_ti_ is the momentary-level residual capturing the unexplained variability for individual *i* at time *t*. At level-2, the intercept of level-1 (β_0_) was defined: γ_00_ is the overall mean or the intercept, γ_01_ to γ_08_ are the between-person associations between the outcomes and time-invariant covariates, and u_0i_ is the person-specific residual deviations or random intercepts for the four outcomes (in respective models). The equation, β_(1-4)i_ = γ_(1–4)0_, means that no random slope effect was added for β_(1-4)i_, with assumptions that the relationships between the time-varying predictors and the outcomes were consistent across individuals, primarily due to the limited sample size.

## Results

### Participant demographics

Out of the 23 participants in the study, 19 participants were included in the analysis due to missingness from the remaining 4 participants. The demographic information of the included (n=19) and excluded (n=4) participants is reported in Table 1. The mean age of the participants was 47.37 years (*SD*=11.58). Majority of them were Caucasian (*n*=16, 84.21%), females (*n*=17, 89.47%), and had completed a bachelor’s degree (e.g., BA, BS) (*n*=9, 47.37%).

**Table 1.**
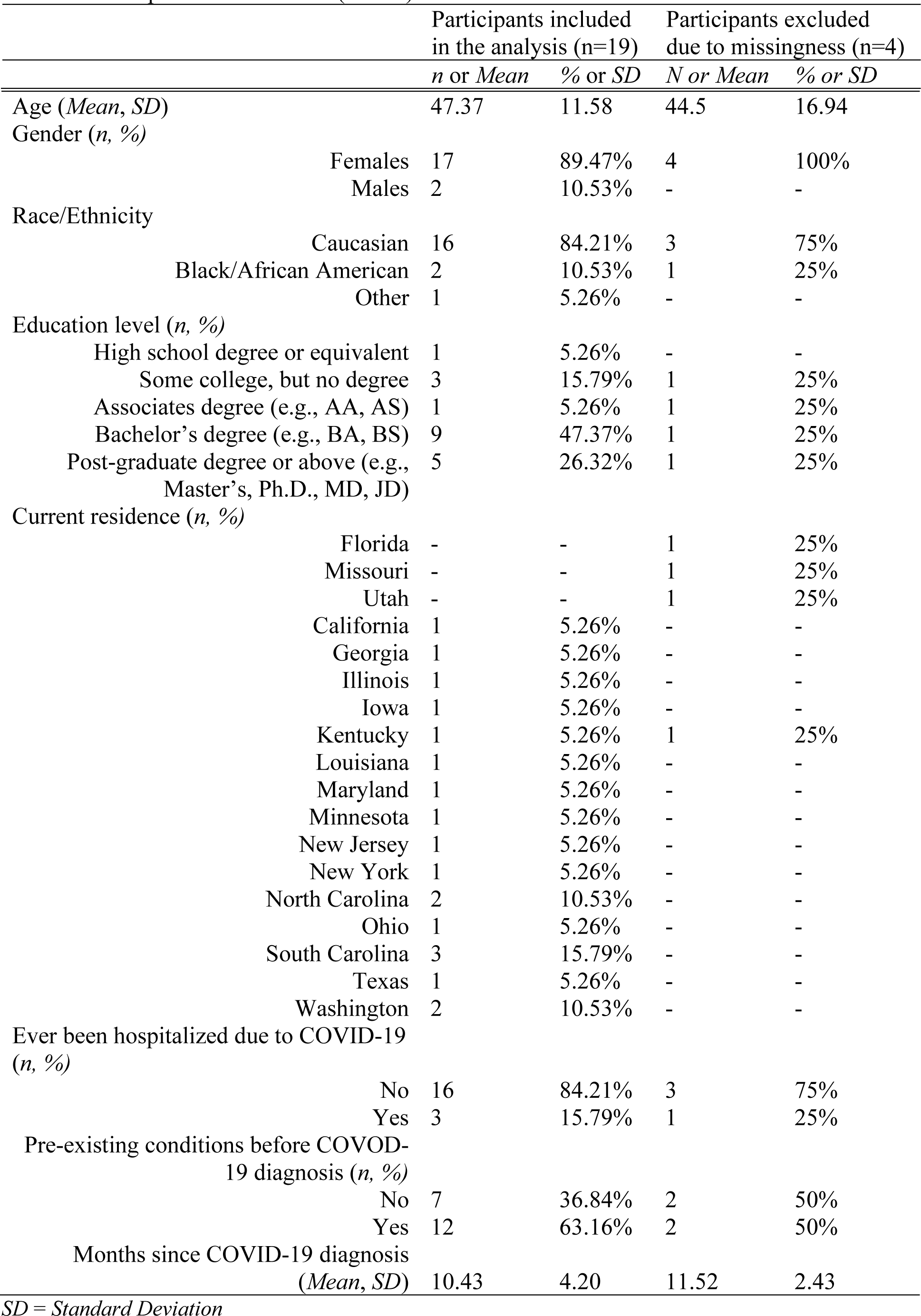
Participant characteristics (*n* = 23)

| Participants included in the analysis (n=19) |  | Participants excluded due to missingness (n=4) |  |
| --- | --- | --- | --- |
| n or Mean | % or SD | N or Mean | % or SD |

|  |  |  |  |  |
| --- | --- | --- | --- | --- |
| Age (*Mean, SD*) | 47.37 | 11.58 | 44.5 | 16.94 |
| Gender (*n, %*) |  |  |  |  |
| Females | 17 | 89.47% | 4 | 100% |
| Males | 2 | 10.53% | - | - |
| Race/Ethnicity |  |  |  |  |
| Caucasian | 16 | 84.21% | 3 | 75% |
| Black/African American | 2 | 10.53% | 1 | 25% |
| Other | 1 | 5.26% | - | - |
| Education level (*n, %*) |  |  |  |  |
| High school degree or equivalent | 1 | 5.26% | - | - |
| Some college, but no degree | 3 | 15.79% | 1 | 25% |
| Associates degree (e.g., AA, AS) | 1 | 5.26% | 1 | 25% |
| Bachelor's degree (e.g., BA, BS) | 9 | 47.37% | 1 | 25% |
| Post-graduate degree or above (e.g., Master's, Ph.D., MD, JD) | 5 | 26.32% | 1 | 25% |
| Current residence (*n, %*) |  |  |  |  |
| Florida | - | - | 1 | 25% |
| Missouri | - | - | 1 | 25% |
| Utah | - | - | 1 | 25% |
| California | 1 | 5.26% | - | - |
| Georgia | 1 | 5.26% | - | - |
| Illinois | 1 | 5.26% | - | - |
| Iowa | 1 | 5.26% | - | - |
| Kentucky | 1 | 5.26% | 1 | 25% |
| Louisiana | 1 | 5.26% | - | - |
| Maryland | 1 | 5.26% | - | - |
| Minnesota | 1 | 5.26% | - | - |
| New Jersey | 1 | 5.26% | - | - |
| New York | 1 | 5.26% | - | - |
| North Carolina | 2 | 10.53% | - | - |
| Ohio | 1 | 5.26% | - | - |
| South Carolina | 3 | 15.79% | - | - |
| Texas | 1 | 5.26% | - | - |
| Washington | 2 | 10.53% | - | - |
| Ever been hospitalized due to COVID-19 (*n, %*) |  |  |  |  |
| No | 16 | 84.21% | 3 | 75% |
| Yes | 3 | 15.79% | 1 | 25% |
| Pre-existing conditions before COVID-19 diagnosis (*n, %*) |  |  |  |  |
| No | 7 | 36.84% | 2 | 50% |
| Yes | 12 | 63.16% | 2 | 50% |
| Months since COVID-19 diagnosis (*Mean, SD*) | 10.43 | 4.20 | 11.52 | 2.43 |

*SD = Standard Deviation*

### Quantitative results

#### Feasibility and acceptability

Mean daily surveys answered were 15.9 (*SD*=9.2). Given that the surveys were sent out for 28 days, the response rate of the participants ranged from 7.14% to 96.43%, with a mean of 57% (*SD*=32.85). Participants with lower response rate were also included in the analysis because even single-day data was crucial to assess the day-to-day impact of MW on mental health outcomes. The total number of days when participants practiced MW ranged from 0 to 24, with a mean of 7.29 days (*SD*=5.25). On overage, the participants practiced MW for 1.72 days (*SD*=1.38) within a week.

Paradata showed that the participants were satisfied with the program in terms of mindfulness skill enhancement (*n*=11) (i.e., attention, mindful breathing, slow walking), pain reduction (*n*=1), psychological well-being promotion (*n*=2) (i.e., emotion regulation and stress relief), and willing to participate in similar programs (*n*=2) (Appendix 1). On a scale of 1-7, the mean scores of key USE variables were: usefulness 4.57 (*SD*=2.02), ease of use 5.04 (*SD*=1.57), ease of learning 5.44 (*SD*=1.87), and satisfaction 4.69 (*SD*=1.43). Higher scores indicate a higher performance on that variable. The overall feasibility, i.e., the mean of mean scores on all four variables was 4.93 (*SD*=1.88).

#### Preliminary efficacy

Table 2 shows the descriptive statistics of primary variables and time-varying covariates. A total of 303 responses were collected. On a scale of 1–7, participants reported moderate levels of positive affect (*M*=3.95, *SD*=1.14), lower levels of negative affect (*M*=3.33, *SD*=1.26), higher level of perceived cognition (*M*=4.38, *SD*=1.3) and mindfulness (*M*=5.29, *SD*=1.17). On average, the participants engaged in 19.86 minutes of physical activity on any given day (*SD*=27.56) beyond doing the MW (if any that day) and practiced MW for 7.29 days (*SD*=5.25). Most of the times when participants responded to the survey, they had not practiced MW up to that time of the day (*n*=198, 65.56%). Participants engaged in all session types, with a higher occurrence of session 3 and 4, and session 5 and 6 (*n*=27, 25.71%). The majority of the participants responded between 8-10 PM (*n*=261, 86.14 %).

**Table 2.** Descriptive statistics for main variables and time-varying covariates (N=19, 303 rows).

|  | n or Mean | % or SD |
| --- | --- | --- |
| Main Variables (*Mean, SD*) |  |  |
| Positive Affect (3 items, scale 1-7) | 3.95 | 1.14 |
| Negative Affect (3 items, scale 1-7) | 3.33 | 1.26 |
| Perceived Cognition (2 items, scale 1-7) | 4.38 | 1.3 |
| Mindfulness (2 items, scale 1-7) | 5.29 | 1.17 |
| Physical activity mins on a given day | 19.86 | 27.56 |
| Number of MW days (*Mean, SD*) | 7.29 | 5.25 |
| MW on a given day (*n, %*) |  |  |
| No | 198 | 65.56% |
| Yes | 104 | 34.44% |
| MW Session Practiced (*n, %*) |  |  |
| Session 1 (week 1) | 18 | 17.14% |
| Session 2 (week 1) | 17 | 16.19% |
| Session 3 and 4 (week 2) | 27 | 25.71% |
| Session 5 and 6 (week 3) | 27 | 25.71% |
| Session 7 and 8 (week 4) | 16 | 15.24% |
| Time of finishing the daily survey (*n, %*) |  |  |
| Before 8 AM | 9 | 2.97% |
| 08-10 AM | 8 | 2.64% |
| 10-12 PM | 8 | 2.64% |
| 12-02 PM | 10 | 3.3% |
| 02-04 PM | 5 | 1.65% |
| 04-06 PM | 2 | 0.66% |
| 06-08 PM | - | - |
| 08-10 PM | 261 | 86.14% |
| 10-12 Midnight | 9 | 2.97% |
| Typical day (*n, %*) |  |  |
| No | 186 | 61.39% |
| Yes | 117 | 38.61% |
| Social Engagement (Number of people) (*Mean, SD*) | 10.15 | 12.59 |

Table 3 presents the results of a multilevel model analysis. The dependent variables included mindfulness, positive affect, negative affect, perceived cognition, and physical activity. The independent variables included practicing MW on a given day or not and the total number of MW days. The models also controlled for day-level and demographic covariates. The preliminary findings on efficacy of the intervention were as follows: (1) MW uptake on a given day was significantly associated with higher positive affect (*β*=0.89, *p*<0.01), lower negative affect (*β*=-0.83, *p*<0.01), higher perceived cognition (*β*=0.52, *p*<0.05), and higher physical activity levels (*β*=0.41, *p*<0.05). (2) Total number of MW days across the study period were significantly and positively associated with higher mindfulness levels (*β*=0.3 *p*<0.01). All models controlled for age, gender, race, education, months since COVID diagnosis, hospitalization due to COVID, pre-existing medical conditions, time of finishing the survey, typical day, and social engagement levels on that day.

**Table 3.** Multilevel Model Results (N=19, 303 rows)

|  | Positive Affect |  | Negative Affect |  | Perceived Cognition |  | Mindfulness |  | Physical Activity |  |
| --- | --- | --- | --- | --- | --- | --- | --- | --- | --- | --- |
|  | *Value* | *SE* | *Value* | *SE* | *Value* | *SE* | *Value* | *SE* | *Value* | *SE* |
| *Fixed Effects* |  |  |  |  |  |  |  |  |  |  |
| (Intercept) | 9.3*** | 2.02 | 10.1*** | 2.52 | 5.79** | 1.78 | 3.63* | 1.55 | 0.53 | 1.03 |
| MW on a given day<br>(No=0, Yes=1) | 0.89** | 0.32 | -0.83** | 0.31 | 0.52* | 0.25 | 0.03 | 0.19 | 0.41* | 0.16 |
| Total MW days | 0.13 | 0.12 | -0.08 | 0.15 | 0.04 | 0.1 | 0.3** | 0.09 | 0.06 | 0.06 |
| Age | 0.03 | 0.04 | -0.14* | 0.05 | 0.02 | 0.04 | -0.04 | 0.03 | -0.02 | 0.02 |
| Gender (Males)<br>Reference: Females | -4.79* | 1.8 | 1.34 | 2.32 | -1.79 | 1.61 | -2.51 | 1.42 | -0.34 | 0.92 |
| Race (Whites)<br>Reference: Others | 1.29 | 1.31 | 0.77 | 1.67 | 0.94 | 1.17 | 2.71* | 1.02 | -0.72 | 0.67 |
| Education | 0.38 | 0.49 | -1.2 | 0.62 | 0.52 | 0.44 | 1.11* | 0.38 | 0.35 | 0.25 |
| Months since COVID diagnosis | -0.23 | 0.16 | 0.42 | 0.2 | -0.07 | 0.14 | -0.23 | 0.13 | -0.09 | 0.08 |
| COVID hospitalization (No=0, Yes=1) | 1.08 | 1.56 | 0.1 | 2.02 | 1.06 | 1.4 | -0.08 | 1.23 | 1.43 | 0.8 |
| Pre-existing medical conditions (No=0, Yes=1) | 4.32** | 1.29 | -3.51 | 1.68 | 2.09 | 1.16 | 3.3** | 1.03 | 1.42 | 0.65 |
| Time of finishing the survey | -0.08* | 0.03 | 0.06 | 0.03 | -0.02 | 0.03 | 0.01 | 0.02 | 0 | 0.02 |
| Typical day (No=1, Yes=0) | -2.61*** | 0.37 | 2.57*** | 0.36 | -1.73*** | 0.29 | -0.24 | 0.23 | -0.22 | 0.19 |
| Social engagement (No=0, Yes=1) | 0.02 | 0.01 | 0.01 | 0.01 | 0.02* | 0.01 | 0.01 | 0.01 | 0.02** | 0.01 |
| *Random Effects* |  |  |  |  |  |  |  |  |  |  |
| Intercept variance |  | 2.76 |  | 5.06 |  | 2.31 |  | 1.88 |  | 0.72 |
| Residual variance |  | 5.48 |  | 5.11 |  | 3.46 |  | 2.04 |  | 1.39 |

SE= Standard Error
\*\*\*p < .001; \*\*p < .01; \*p < .05; † < .1

### Qualitative results

Analysis of the Paradata identified two main themes: (1) positive influences of the intervention program and (2) challenges arising in the intervention program. The structure of the themes and dimensions are presented in Appendix 1.

#### Positive outcomes of the intervention program

Most posts were identified as messages or dialogs about the benefits of participating in the intervention program. The benefits included “awareness/mindful enhancement”, “physical enhancement”, and “emotional change”. In terms of “awareness/mindful enhancement”, participants noted that the MW exercises promoted their experiences to focus on being present in the moment and be aware of their bodily changes in several aspects. The changes included “physiological awareness” (e.g., “*I am finding it easier to mindfully walk while paying attention to my breath*”) (*n*=7; 32%), “behavioral awareness” (e.g., “*I realized walking one step per second was actually difficult*”) (*n*=9; 39%), and “self-consciousness/focus on the self” (e.g., “*Now I notice that when people walk by me, I’m still focusing on my steps*”) (*n*=2; 7%). Participants noted that they experienced physical enhancement during the intervention. The enhancement involves “COVID-19 symptom relief” (e.g., “*I was unaware that I was walking one step per second with my post COVID symptoms…During the session, I wasn’t concentrating on it—I suppose I knew it [COVID symptoms] was there, but the session distracted me from it”)* (*n*=1; 4%) and “physical comfort” (e.g., “*Now I feel more comfortable*”) (*n*=1; 4%). One participant mentioned an emotional change, noting that he/she felt pleasant when practicing MW (e.g., “*While walking this evening, I notice the smell of fresh cut grass. Haven’t been something that I’ve really paid much attention to. It was refreshing*”) (*n*=1; 4%).

#### Challenges arising in the intervention program

Participants mentioned several challenges arising in the intervention program. Some challenges were related to environmental factors, such as “bad weather” (e.g., “*It was bad weather that day…I got overwhelmed by paying attention to my breathing*.”) (*n*=2; 7%) and “flooding” (e.g., “*Unfortunately, my neighborhood is flooded, and I will be walking here this week”*) (*n*=1; 4%). Participants also noted that their engagement was influenced by factors derived from the intervention program, including “the difficulty of learning MW skills” (e.g., “*I thought this last exercise was more difficult than the others. I felt like I was more focused on getting the skills correct than just being present and in the moment.*”) and “negative emotion arising from MW practice” (e.g., “*Truthfully, I got overwhelmed by paying attention to my breathing. It felt like I had covid again.*”) (*n*=1; 4%).

## Discussion

### Principal findings

This current study tested the feasibility and preliminary efficacy of digitally delivering the MW intervention to long haulers via social media platform, Facebook, using a mixed methods design including daily diary surveys. Our findings demonstrate strong feasibility in terms of *adherence*, including low dropout rates, reasonable response rate to the daily diary surveys, and high engagement with MW weekly and throughout the study period; and *acceptability*, with higher-than-average score on the USE scale (4.9/7), demonstrating strong usefulness, ease of use, satisfaction, and ease of learning. In terms of preliminary efficacy, the multilevel analysis on the daily diary surveys indicated that on days participants engaged in MW, they reported increased positive affect, perceived cognition, and physical activity levels, and reduced negative affect. Additionally, the total number of days of MW practice across the study period was linked to enhanced mindfulness on a given day.

The quantitative findings on preliminary efficacy were supported by the qualitative findings as participants reported improved physical comfort through symptom relief, feeling pleasant when practicing MW, and physiological-, behavioral-, and overall self-awareness. Findings on feasibility were enriched through understanding two broad types of reasons for missing the MW sessions, including external factors such as bad weather and flooding, and difficulties in learning MW skills. Overall, participants reported not only improved mental well-being but also witnessed an increase in physical comfort and physical activity levels on a given day.

### High acceptability

The acceptability of our MW program was assessed through dropout rates, responsiveness to daily surveys, and engagement with the MW sessions. All 23 participants who joined the Facebook group committed to the 4-week MW program, with only 4 showing missingness in daily diary surveys. The sustained participation of most participants in the MW program, as evidenced by their ongoing involvement, indicated a strong interest in such interventions among long haulers. Participants responded to daily surveys about 60% of the time (SD=33), similar to previous EMA research with long haulers (Dreimann, 2021; Junge, 2023), demonstrating substantial commitment to the intervention’s daily self-monitoring.

In MW sessions, participants showed maximum engagement with sessions 3 and 4 (week 2) (n=27, 25.71%), and sessions 5 and 6 (week 3) (n=27, 25.71%). Each of these sessions included 10 minutes of mindful breathing, with sessions 5 and 6 including an additional 10 minutes of step focused. The lowest engagement occurred in sessions 7 and 8, which included an additional full body scan (n=16, 15.24%). This was also indicated by a participant in the qualitative surveys, who reported difficulty in learning the “last exercise”. Similarly, session 1 (n=18, 17.14%) and session 2 (n=16, 16.19%), which included no mindful breathing and only 5 minutes of mindful breathing, respectively, had lower engagement. This suggests a preference for a balanced approach of mindful breathing and step-focused walking during the 30 minutes slow walk among long haulers.

The sessions were released at the start of the week, with participants encouraged to practice each session at least once, aiming for two sessions per week. For instance, in week 1, participants were expected to practice sessions 1 and 2. Given that, participants engaged in MW for 1.72 days per week (SD=1.38), roughly aligning with the expectation of two sessions weekly. Over the study period, participants practiced MW for an average of 7.29 days (SD=5.25), close to the anticipated 8 sessions throughout the study.

The average level of acceptability, measured using the USE scale (4.9/7), demonstrated strong usefulness, ease of use, satisfaction, and ease of learning. Some participants, however, reported difficulties in learning mindfulness exercises, especially the full body scan in the last exercise. Our results highly complement the previous MW intervention studies, where older adults (Yang & Conroy, 2018), breast cancer patients (Schröder et al., 2022), and community residents (Jones et al., 2021) have reported high acceptability to the MW intervention. Despite promising results, the various engagement across sessions suggest a need for personalized motivational strategies and addressing potential physical limitations to enhance future program engagement (Aggarwal et al., 2023).

### Improved Mental Well-being and Physical Activity Levels

Our MW intervention demonstrated an improvement in participants’ mental well-being, including heightened positive affect, perceived cognition, mindfulness, and reduced negative affect, as well as an increase in participants’ daily physical activity levels beyond the duration of MW. These findings are aligned with previous MW intervention research that demonstrated similar findings with older adults (Yang et al., 2021; Yang & Conroy, 2018), breast cancer survivors (Schröder et al., 2022), college students (Ma et al., 2023), psychologically distressed individuals (Teut et al., 2013), general adults (Gotink et al., 2016), young adults (Bigliassi et al., 2020), and among adults with inadequate physical activity levels (Shi et al., 2019). The Psychoneuroimmunological Framework offers insights into the observed outcomes. MW induces a relaxation response, reducing chronic inflammation in long haulers (Magan & Yadav, 2022), promotes attention regulation, countering cognitive challenges like ‘brain fog’ (Jha et al., 2007), fosters body awareness and interoception, aiding in symptom management (Mehling et al., 2012), releases endorphins that help in pain relief and mood elevation (Boecker et al., 2008), and promotes emotion regulation and neuroplasticity, leading to improved emotional resilience (Taren et al., 2017).

Similarly, our findings on increased physical activity are also supported by previous MW intervention research. Breast cancer patients reported increased physical exercise (Schröder et al., 2022), adults with inadequate activity exhibited significant improvements in physical activity (Shi et al., 2019), and patients with chronic obstructive pulmonary disease also reported improved the exercise capacity (Lin & Yeh, 2021). This preliminary finding suggests that MW could be a viable approach for improving physical activity in long haulers. The low-intensity nature of MW makes it an accessible exercise, aiding in rebuilding physical strength and endurance without excessive exertion (Lin & Yeh, 2021; Sun et al., 2022; Teut et al., 2013). Our study uniquely contributes to the existing literature by exploring the mental and physical health benefits of digitally delivered MW interventions with long haulers. The encouraging outcomes strongly advocate for the adoption of MW interventions in managing chronic conditions such as long COVID.

### The Need for Online Interventions

Our program uniquely harnessed the power of social media, executing every phase from recruitment to evaluation. With the increasing penetration of smartphones and internet access, even in developing countries, such interventions hold promise for wide-reaching impact, making quality care accessible to broader populations (George et al., 2012). Given our findings on the feasibility of delivering MW intervention via a social media platform, future studies should aim to test the scalability and affordability of such online interventions, especially in low-income settings. For instance, a study could be designed to target long-haulers in the rural communities of sub-Saharan Africa, where, despite the challenges of infrastructure, mobile connectivity has witnessed a significant surge (GSMA, 2023; Silver et al., 2019). Such studies would offer insights into potential barriers and facilitators specific to these settings, like local digital literacy levels (Sharma et al., 2016) or cultural perceptions of online health interventions (X. Liu et al., 2010). Moreover, studies should also consider integrating vernacular languages and culturally relevant content to enhance engagement and relatability (X. Liu et al., 2010). Assessing the cost-effectiveness of these interventions in such settings, where healthcare resources are often limited, will be vital (Aggarwal et al., 2023; Mudiyanselage et al., 2023).

### Social Engagement as a Catalyst

While MW is typically a solo activity, executing our MW program through a Facebook group leveraged social engagement. This platform’s social nature created a sense of community, with participants actively engaging through comments and posts. They shared experiences and motivated each other, exemplifying the therapeutic potential of online social groups for mutual support in pain and recovery (Williams et al., 2009). Watching peers’ progress motivated many, reinforcing their commitment and fostering a sense of belonging, crucial for emotional well-being, especially for long haulers who often feel isolated (Pfefferbaum & North, 2020; Umberson & Montez, 2010). Thus, the program extended beyond physical and mental benefits to socio-emotional gains through shared experiences.

Future research should explore the impact of online social groups, particularly in collectivist societies where community and social connections ties are vital (Triandis & Gelfand, 2012).. Therefore, online platforms that emphasize collective healing, shared experiences, and mutual support can be particularly impactful. Future studies could investigate how tailored interventions for these societies can leverage cultural values and social structures to enhance the therapeutic potential of online groups. Understanding virtual community dynamics in these societies could lead to creating culturally relevant, effective interventions that can address the unique needs of individuals in variety collectivist cultures (Harrell, 2018).

### Strengths and Limitations

A key strength of our study was the use of social media from recruitment to evaluation, especially beneficial during the COVID-19 pandemic for safe, remote participation. For quantitative assessments, our study used the daily diary surveys, which offered dynamic monitoring of the intervention and nuanced insights into its day-to-day impact, providing a granular understanding often missed in retrospective assessments (Bolger, 2013). Moreover, the extraction of paradata from the Facebook group added a rich layer to our findings. Paradata, in this context, captured the lived experiences of participants, their spontaneous reactions, and organic discussions, providing an unfiltered perspective on the intervention’s effects and areas of improvement (Couper, 2017).

The study, while promising, presents several limitations. The participants were mainly Caucasian and female with high education attainment, limiting the generalizability of our findings. The recruitment exclusively from Facebook groups might introduce selection bias, representing a subset of long haulers more digitally active or inclined to join such groups. The collection of only daily survey data restricts our understanding of the intervention’s prolonged effects, future studies should include extended follow-ups. The absence of a control group in the study design limits our ability to definitively attribute observed outcomes to the intervention. While Paradata from Facebook groups offers valuable insights, it might not capture the full range of participants’ experiences. Future studies need to conduct in-depth interviews to gather more reliable information.

## Conclusion

Our study demonstrated feasibility and preliminary efficacy of a digitally delivered MW program through Facebook for long haulers. Positive results on adherence and acceptability reflect the suitability of digitally delivering MW intervention. Participants reported enhanced positive affect, perceived cognition, and physical activity, along with reduced negative affect on days they engaged in MW. Additionally, the total number of MW practice days was linked to mindfulness on a given day. Qualitative findings complemented these results as participants shared their experiences of symptom management, feeling pleasant, and increased awareness through MW. Conclusively, digitally delivered MW could be a viable and effective approach for not only improving mental well-being but also physical activity in long haulers. The encouraging outcomes strongly advocate for the adoption of MW interventions in managing chronic conditions such as long COVID. Especially given the accessibility and low resource demands of online delivery, this intervention has the potential to be particularly beneficial in contexts where traditional healthcare delivery faces challenges.

## Data Availability

Data Availability Statement: The data analyzed during the current study is available on request to the corresponding author.

---

**Appendix S1.**
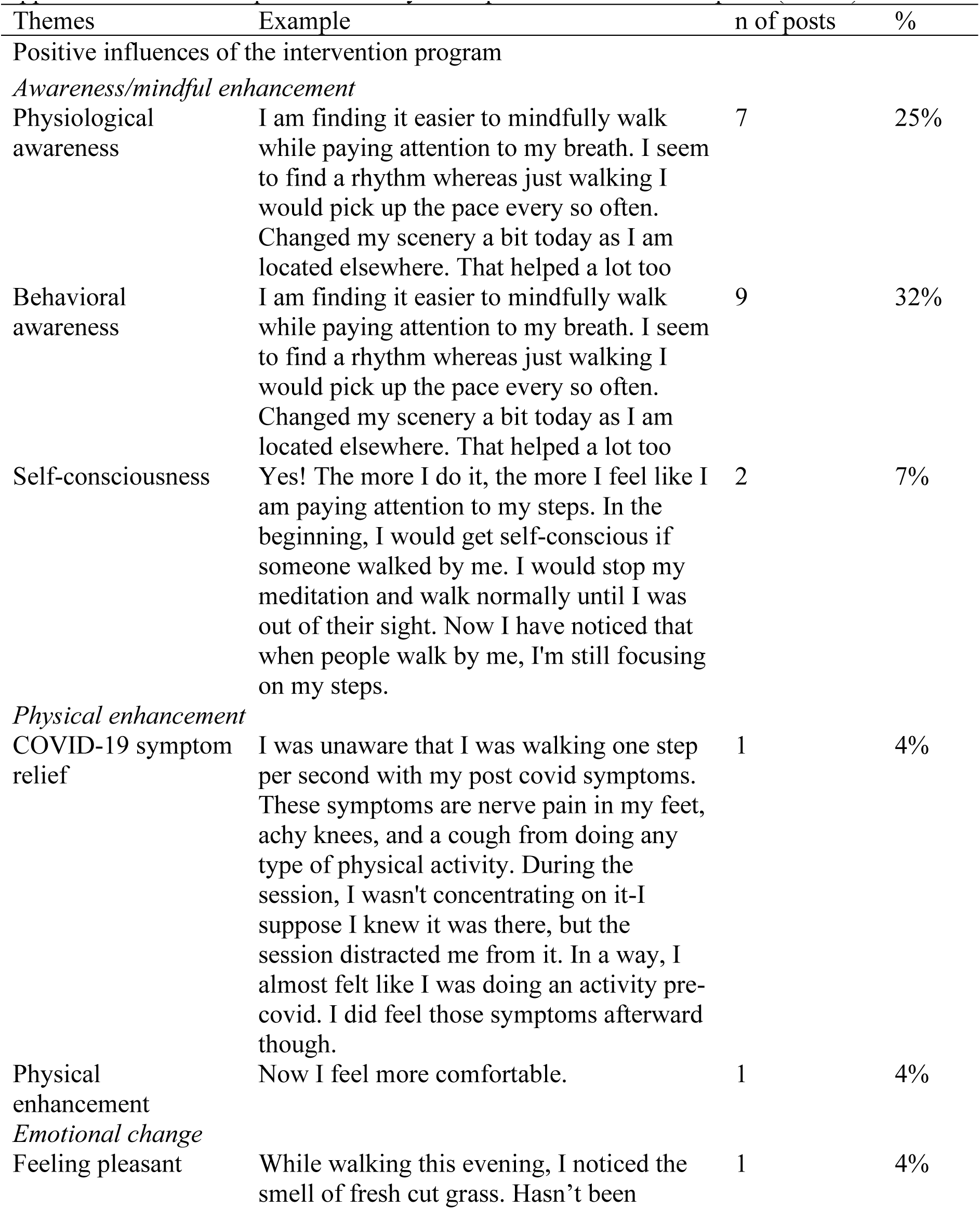
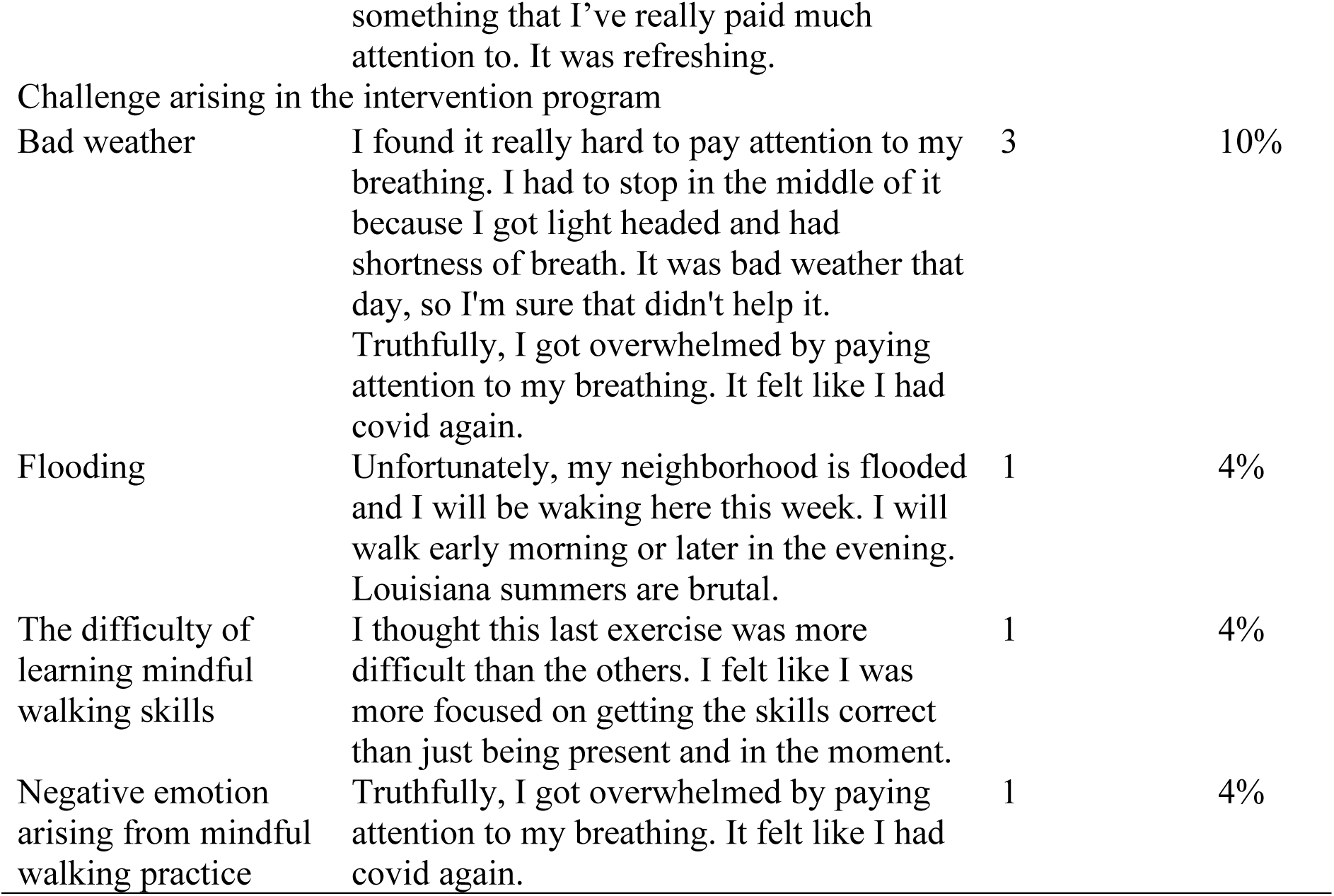
Results of qualitative analysis on paradata of Facebook posts (*n* = 28)

